# Genomic investigation of multi-species and multi-variant blaNDM outbreak reveals key role of horizontal IncN and IncX3 plasmid transfer

**DOI:** 10.1101/2023.08.02.23293478

**Authors:** Nenad Macesic, Adelaide Dennis, Jane Hawkey, Ben Vezina, Jessica A. Wisniewski, Hugh Cottingham, Luke V. Blakeway, Taylor Harshegyi, Katherine Pragastis, Gnei Zweena Badoordeen, Pauline Bass, Andrew J. Stewardson, Amanda Dennison, Denis W. Spelman, Adam W.J. Jenney, Anton Y. Peleg

## Abstract

**Objectives:** New Delhi metallo-beta-lactamases (NDMs) are major contributors to the spread of carbapenem resistance globally. In Australia, NDMs were previously associated with international travel but from 2019 we noted increasing NDM episodes. We conducted an investigation to determine the clinical and genomic epidemiology of NDM-carriage at a tertiary Australian hospital from 2016-2021.

**Methods:** We identified 49 patients with 84 NDM-carrying isolates in an institutional database and collected clinical data from electronic medical records. Short- and long-read whole genome sequencing was performed on all isolates. Completed genome assemblies were used to assess the genetic setting of *bla*_NDM_ genes and compare NDM plasmids.

**Results:** Of 49 patients, 38 (78%) were identified in 2019-2021 and only 11/38 (29%) reported prior travel compared with 9/11 (82%) in 2016-2018 (*P=*0.037). In patients with NDM infection, crude 7-day mortality was 0% and 30-day mortality was 14% (2/14 patients). NDMs were noted in 41 bacterial strains (i.e. species/sequence type combinations). Four NDM variants (*bla*_NDM-1_, *bla*_NDM-4_, *bla*_NDM-5_, *bla*_NDM-7_) were detected across 13 plasmid groups. We noted a change from a diverse NDM plasmid repertoire in 2016-2018 to the emergence of conserved *bla*_NDM-1_ IncN and *bla*_NDM-7_ IncX3 epidemic plasmids with inter-strain spread in 2019-2021. These plasmids were noted in 19/38 (50%) patients and 35/68 (51%) genomes in 2019-2021.

**Conclusions:** Increased NDM case numbers were due to local circulation of two epidemic plasmids with extensive inter-strain transfer. Our study underscores the challenges of outbreak detection when horizontal transmission of plasmids is the primary mode of spread.

## Introduction

Carbapenem-resistant Gram-negative bacteria cause significant morbidity and mortality globally and were declared a critical priority by the World Health Organization [1, 2]. New Delhi metallo-beta-lactamases (NDMs) are significant contributors to the spread of carbapenem resistance [3]. While endemic in South and Southeast Asia, NDMs are increasingly noted in other settings (e.g. North America and Europe), where they have caused substantial clonal outbreaks [4, 5]. There is a paucity of treatment options for metallo-beta-lactamase infections [6, 7]. Clinical data for new treatments such as cefiderocol and aztreonam-avibactam combinations are limited, while older agents such as polymyxins have significant toxicities [6, 7]. This makes stopping NDM spread an urgent priority.

In Australia, NDMs have traditionally been associated with travel from endemic countries [8]. From 2019 we noted increasing numbers of NDM episodes at our institution, which persisted despite COVID-19 travel restrictions. These episodes involved multiple bacterial host species, leading us to hypothesise that local NDM spread may be occurring through horizontal transfer of NDM plasmids. We therefore aimed to determine the clinical and genomic epidemiology of NDM carriage at our institution from 2016 to 2021. More specifically, we sought to apply long-read sequencing approaches to characterise NDM genetic settings accurately and understand within-patient diversity and transfer dynamics of NDM plasmids.

## Methods

### Study setting and population

The study was approved by the Alfred Hospital Ethics Committee. We reviewed an institutional database of carbapenem-resistant Gram-negative isolates spanning 2002-2021 at a healthcare system comprising a tertiary hospital, a community hospital, and a geriatric/rehabilitation hospital. We identified NDM carriage in 49 patients (84 isolates) from September 2016-June 2021. Clinical data were extracted from the electronic medical record (including patient movement data) and a Charlson co-morbidity index score calculated. NDM isolates were classified as associated with colonisation or infection [9]. Crude mortality at 7- and 30-days was recorded. We divided the study into two equal periods from first to final NDM patient (2016-2018 and 2019-2021). Patients were screened for carbapenemase-producing organisms following discharge from the intensive care unit (ICU), if they had a history of international hospitalisation, or were contacts of patients with carbapenemase-producing organisms, per state guidelines [10]. Due to absence of systematic NDM surveillance, we considered that patients may have been colonised in the 30 days prior to first isolation of NDM-carrying organism and identified overlaps on the same ward at the same time as potential between-patient transmission events.

### Isolate selection and genomic analyses

Routine antimicrobial susceptibility testing was performed using Vitek2 (BioMérieux) to presumptively identify NDM-carrying isolates. NDM presence was confirmed using the Xpert Carba-R multiplex PCR assay (Cepheid). All 84 NDM-carrying isolates identified were included in the study and underwent short-read (Illumina) and long-read (Oxford Nanopore Technologies) whole genome sequencing (WGS). One genome was excluded from further analysis due to species mismatch.

Genomic analyses (detailed in Supp. Methods) included resistance gene and plasmid replicon detection with Abricate v.1.0.0 and *in silico* multi-locus sequence typing (MLST) using ‘mlst’ v.2.19.0 [11, 12]. We performed core genome-based phylogenetic analyses on key STs (≥2 genomes available from ≥2 patients) to create a core genome alignment and calculate pairwise single nucleotide variant (SNV) distances. We identified NDM-carrying contigs that were putative plasmids in long-read assemblies using Abricate. MOB-typer v1.4.9 was used to determine plasmid replicons present and identify plasmid groups according to its clustering algorithm [13]. We compared individual plasmids (progressiveMauve v2.4.0.r4736 [14]) and assessed NDM flanking regions (Flanker v0.1.5 [15]).

### Statistical analysis

Categorical variables were compared using χ^2^ or Fisher’s exact tests and continuous variables were compared using the Student’s t-test or Mann-Whitney-Wilcoxon, as appropriate, in R v4.1.1.

## Results

### Study population and clinical characteristics

Of 49 patients with NDM carriage at our institution, 38 (78%) were identified from 2019-2021. Clinical characteristics of patients are shown in Table 1. A history of international travel in the prior year was noted in 20/49 (41%) patients. Significantly fewer patients reported travel in 2019-2021 compared with 2016-2018 (11/38 [29%] vs 9/11 [81%], *P=*0.037). NDM was detected median 5 days (IQR 0-17 days) after admission. Most patients (36/49, 73%) were admitted from the community. 33/49 (67%) patients had a history of hospitalisation in the three months prior including admission to ICU in 15/33 (45%) patients. The first NDM isolate detected was associated with infection in 7/49 (14%) patients. NDM infections developed in 7/42 (17%) patients with initial NDM colonisation (Supp. Table 1). Crude 7- and 30-day mortality in patients with NDM infections were 0% and 14% (2/14 patients), respectively.

**Table 1.** Clinical characteristics of study cohort.

| **Clinical characteristic** |  | **n = 49** |
| --- | --- | --- |
| Period of NDM detection |  |  |
|  | 2016-2018 | 11 (22%) |
|  | 2019-2021 | 38 (78%) |
| Demographics |  |  |
|  | Median age (IQR) | 62 (44 – 74) years |
|  | Male gender | 33 (67%) |
| Comorbidities |  |  |
|  | Pulmonary disease | 14 (29%) |
|  | Diabetes mellitus | 11 (22%) |
|  | Malignancy | 9 (18%) |
|  | Dialysis | 5 (10%) |
|  | Solid organ transplant recipient | 4 (8%) |
|  | Haemopoietic stem cell transplant recipient | 4 (8%) |
| Median Charlson Comorbidity Index Score (IQR) |  | 2 (1 – 3) |
| Risk factors |  |  |
|  | Admitted from community | 36 (73%) |
|  | Admission to hospital in prior three months | 33 (67%) |
|  | Admission to intensive care unit in prior three months | 15 (31%) |
|  | Receipt of antibiotics in prior three months | 39 (80%) |
|  | International travel in prior year | 20 (41%) |
|  | Travel to South Asia | 13 (27%) |
|  | Travel to Southeast Asia | 6 (12%) |
|  | Travel to Middle East | 1 (2%) |
| Initial NDM infection |  | 7 (14%) |
| Initial NDM colonization |  | 42 (86%) |
|  | Subsequent NDM infection | 7 (14%) |

Abbreviations: IQR – Interquartile range; NDM – New Delhi metallo-beta-lactamase

### Isolate characteristics and genomic epidemiology

NDM-carrying bacteria were highly diverse with 41 bacterial strains (defined as unique species/MLST combinations) noted (Figure 1, Supp. Table 2 and Supp. Figure 1) and no dominant strain. Of 12 strains found in ≥2 patients, six strains had between-patient pairwise SNV distances <20 SNVs, suggestive of possible clonal spread (Supp. Table 3). Four NDM variants were detected: *bla*_NDM-1_ (48/83 genomes, 58%), *bla*_NDM-5_ (20/83, 24%), *bla*_NDM-7_ (12/83, 14%) and *bla*_NDM-4_ (3/83, 4%). Non-NDM carbapenemase genes were detected in 11/83 (13%) genomes (Supp. Table 2) and 10/83 (12%) genomes carried *mcr-9*.*1*, a colistin resistance determinant. We noted changes in epidemiology from *E. coli* in 2016-2018 (13/15 [86%] genomes) to non-*E. coli* species in 2019-2021 (54/68 [79%] genomes, *P<*0.001), and from *bla*_NDM-5_ (11/15 [73%] genomes) to non-*bla*_NDM-5_ variants (59/68 genomes [87%], *P<*0.001).

**Figure 1.**
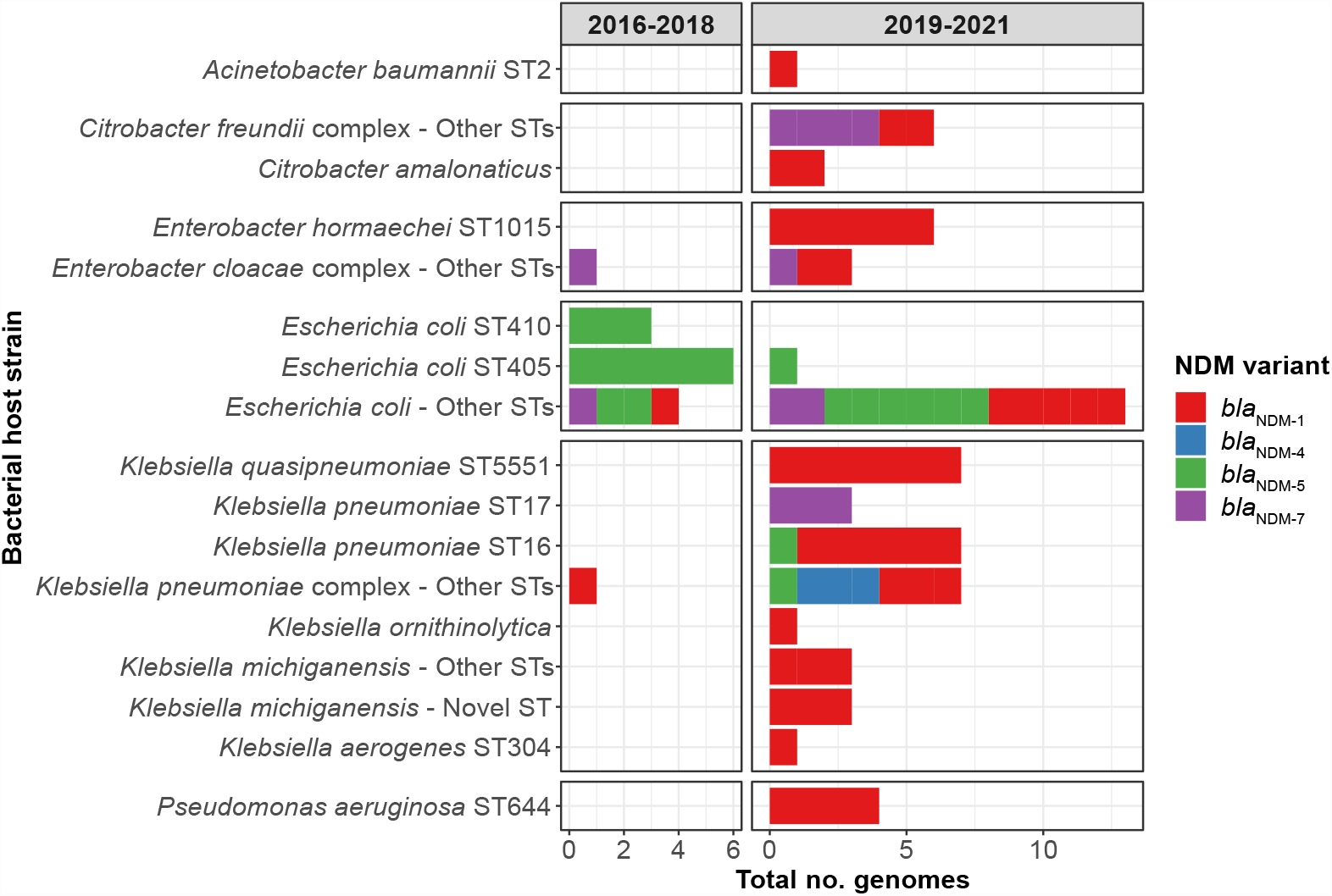
New Delhi metallo-beta-lactamase variants and bacterial host strains 2016-2021. NDM variants and bacterial host strains shown over two study periods. There was a diversity in both the bacterial hosts and NDM variants, with 4 NDM variants being noted across 41 bacterial host strains, with a shift from NDM-5 and *Escherichia coli* in 2016-2018 to NDM-1/NDM-7 and non-*E. coli* species in 2019-2021. Abbreviations: NDM – New Delhi metallo-beta-lactamase; No. – number; ST – sequence type.

### Plasmid analyses

We detected 13 distinct plasmid groups, of which 3/13 had multiple NDM variants present (Figure 2A) and 8/13 were noted in multiple bacterial host strains (Figure 2B). While in 2016-2018 there was a diversity of plasmid groups (8 groups across 11 patients), in 2019-2021 there were three dominant epidemic plasmid groups (*bla*_NDM-1_ IncN, *bla*_NDM-5/NDM-7_ IncX3 and *bla*_NDM-1_ IncC) that were noted in 30/38 (79%) patients and 49/68 (71%) genomes in that period (Figure 2A).

**Figure 2.**
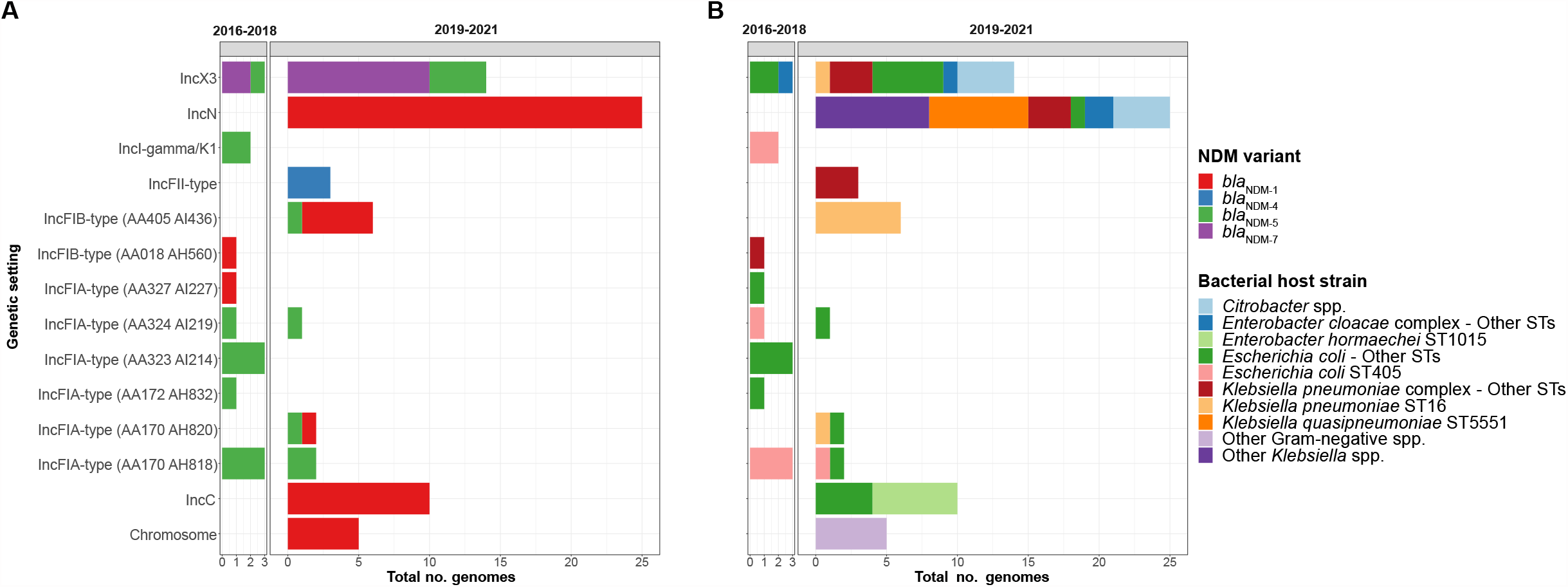
Genetic settings of *bla*_NDM_ 2016-2021. **Fig. 2A:** Genetic settings of *bla*_NDM_ and corresponding NDM variants over study, as defined by two study periods. We detected NDM variants in 13 distinct plasmid groups, as well as integration into the bacterial chromosome. **Fig. 2B:** Genetic settings of *bla*_NDM_ and corresponding bacterial host strains over the study, as defined by two study periods. Abbreviations: NDM – New Delhi metallo-beta-lactamase; No. – number; ST – sequence type.

*bla*_NDM-1_ IncN plasmids were the largest group (25 genomes in 14 patients) and were highly homogenous with 1 SNV across the backbone and minor structural variation near *bla*_NDM-1_ due to a palindromic sequence (Figure 3, Supp. Table 4). IncX3 plasmids were structurally almost identical regardless of NDM variant, but *bla*_NDM-5_ IncX3 plasmids carried 11 SNVs across the backbone compared with 2 SNVs in *bla*_NDM-7_ IncX3 plasmids. Despite belonging to the same plasmid group, IncC plasmids were more diverse with 23 SNVs across the backbone with an obvious sub-cluster of 6 near-identical plasmids from 4 patients (median pairwise SNV distance 1). We also noted mosaic plasmids in all these groups including IncN/IncL/M, IncC/IncHI2A and IncC/IncFIB/IncFII plasmids.

**Figure 3.**
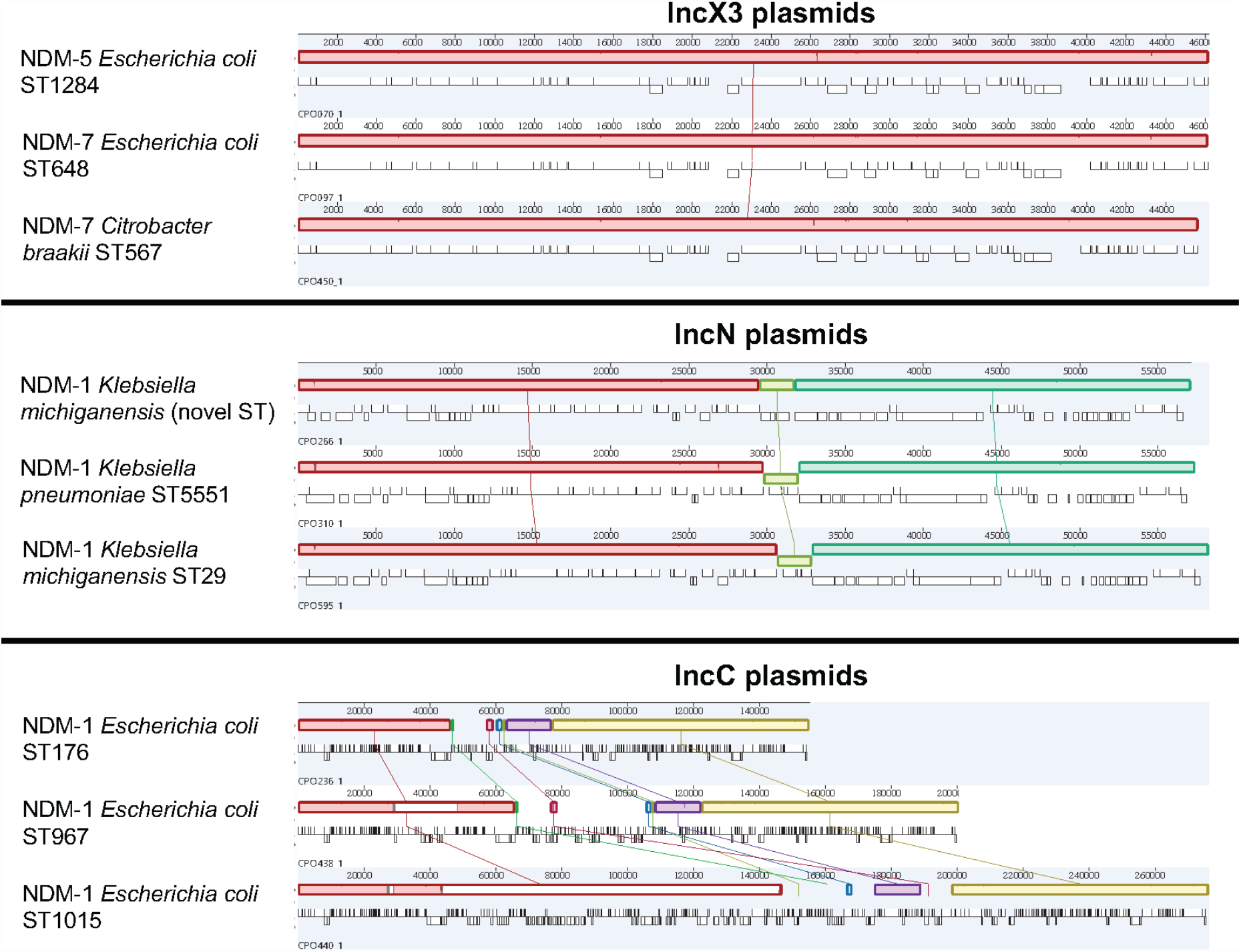
Comparative analyses of NDM IncX3, IncN and IncC plasmids. We aligned representative plasmids from each of the three epidemic NDM plasmid groups from our study. Each coloured field represents a locally collinear block, a homologous region of sequence shared by multiple plasmids without any rearrangement of that region. Same colours indicate the same regions present in different plasmids. IncX3 plasmids were structurally nearly identical regardless of NDM variant. *bla*_NDM-1_ IncN plasmids had minor structural variation near *bla*_NDM-1_ due to a palindromic sequence. While belonging to the same plasmid group, IncC plasmids were more diverse with three plasmid sub-types, as shown. Abbreviations: NDM – New Delhi metallo-beta-lactamase; ST – sequence type.

We compared these epidemic plasmids to plasmids circulating globally (Supp. Figure 2 and Supp. Table 5). IncX3 plasmids were almost identical to global NDM IncX3 plasmids (100% coverage and 99.96% identity). IncN and IncC plasmids were divergent to NDM global plasmids (95% coverage/99.93% identity, 93% coverage/99.99% identity, respectively).

Other plasmid groups with multiple plasmids available for comparison showed larger scale structural differences despite sharing a similar backbone (Supp. Figure 3). Each NDM flanking region was associated with a single plasmid group, except for a transposon capable of carrying two NDM variants (*bla*_NDM-1_ and *bla*_NDM-5_) that was found across seven plasmid groups (Supp. Figure 4). All patients but one with this flanking region had a history of travel to South or Southeast Asia. In one patient we noted both the *bla*_NDM-5_ IncX3 and IncFIA-type (AA170 AH820) plasmids were carrying this flanking region (Supp. Figure 4 - Plasmids 3 and 6, Supp. Figure 5 - Patient 4). This transposon was not found in any other IncX3 plasmids in the study, thus raising the possibility of movement of a transposable element between different plasmids within the patient.

### Within-patient plasmid analysis

Multiple genomes were available for analysis in 18/49 (37%) patients. Six patients had persistent strain colonisation (multiple isolates of the same bacterial strain with same NDM plasmid) and four patients had multiple colonisation events (presence of different plasmid groups) including two patients with plasmids carrying different NDM variants (Supp. Figure 5). In the remaining 8/18 patients, we detected the same plasmid group across different bacterial hosts. We compared plasmids within patients to distinguish potential inter-strain plasmid transfer from multiple colonisation events. The 7/8 patients colonised with *bla*_NDM-1_ IncN and *bla*_NDM-7_ IncX3 epidemic plasmids had highly similar plasmids suggestive of interspecies transfer (Figure 4A and 4B). For the patient colonised with non-epidemic plasmids (Figure 4C), there were significant structural re-arrangements of an IncFII-type plasmid making it less likely that inter-strain plasmid transfer had occurred. In patients with multiple plasmids of the same plasmid group there was a median SNV distance of 0 (range 0 – 267), which is shown by plasmid group in Supp. Table 6.

**Figure 4.**
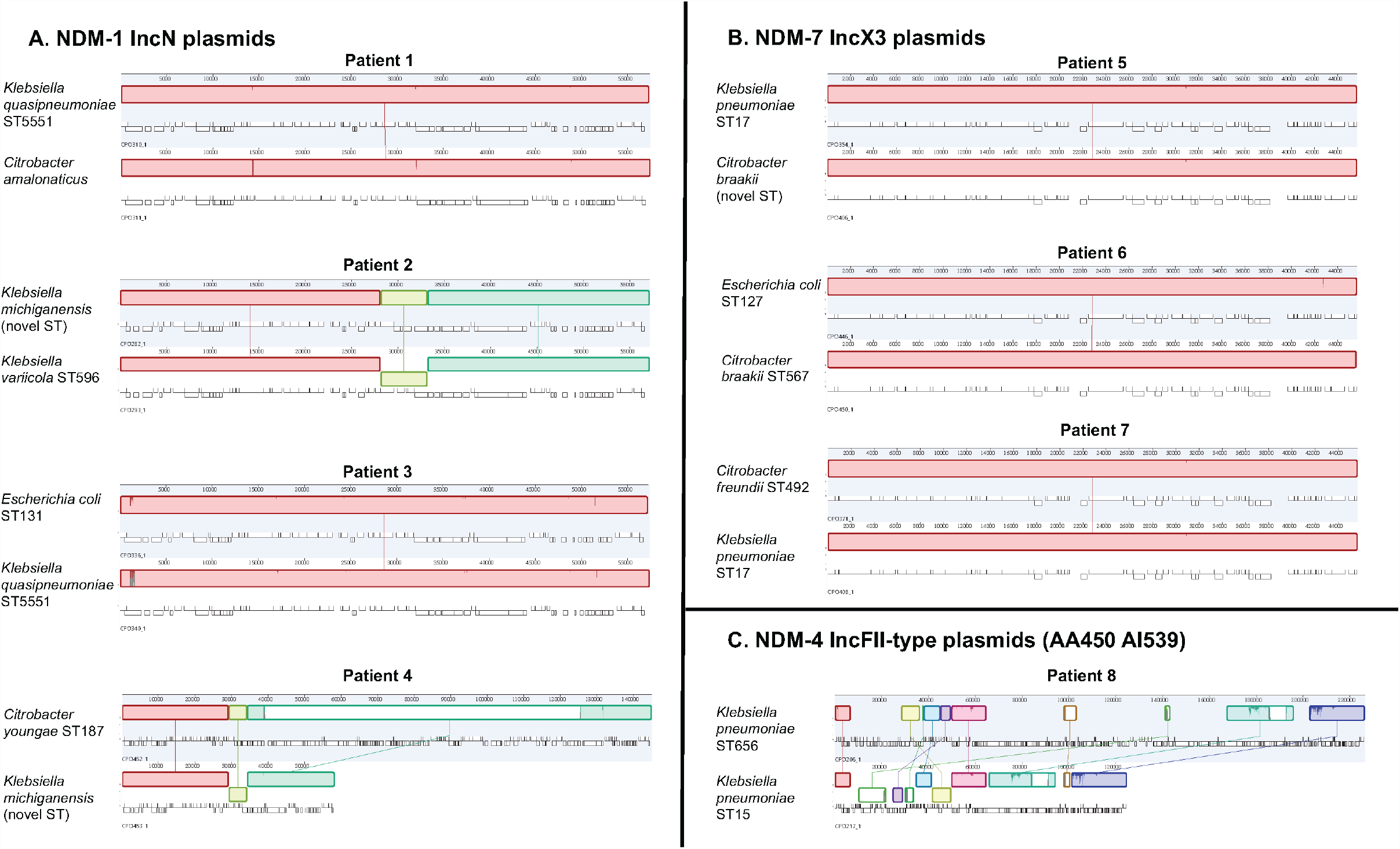
Analysis of potential within-patient plasmid transfer between bacterial host strains. Eight patients were noted to have the same NDM variant/plasmid group combinations across multiple bacterial host strains. Here we aligned plasmids from each of the bacterial host strains, with Fig. 5A showing NDM-1 IncN plasmids, Fig. 5B showing NDM-7 IncX3 plasmids and Fig 5C showing NDM-4 IncFII-type (AA450 AI539) plasmids. The bacterial host strains are shown on the left of the plasmids. Each coloured field represents a locally collinear block, a homologous region of sequence shared by multiple plasmids without any rearrangement of that region. Same colours indicate the same regions present in different plasmids. Abbreviations: NDM – New Delhi metallo-beta-lactamase; ST – sequence type.

### Transmission analysis

We analysed potential in-hospital transmission events by combining detailed patient movement data with plasmid-level genomic data. We defined a potential transmission event when there was spatiotemporal overlap between patients that shared closely related NDM plasmids (same plasmid group and NDM variant with ≤5 SNVs difference in backbone). Despite multiple patients having highly similar plasmids, only 8 patients were linked in 5 potential transmission events, all involving the IncN plasmid (Supp. Figure 6). Despite using criteria aimed at increasing sensitivity, we were unable to directly link 41/49 (84%) patients.

## Discussion

In this study, we combined clinical data with insights from long-read genomic sequencing to analyse the epidemiology of NDM-carrying bacteria at our institution. Although reports of previous NDM outbreaks noted clonal spread of specific strains such as *K. pneumoniae* ST147 and ST307 [5, 16], increased case numbers in our institution were due to circulation of several epidemic plasmids. For most patients, we were unable to document clear healthcare-associated NDM acquisition, highlighting the possibility of acquisition in the community or through undetected sources in the hospital. Ultimately 29% patients developed NDM infection and the 30-day crude mortality rate among these patients was 14%, consistent with previously reported mortality rates of ∼10-20% [17, 18]. Our study also underscored the challenges of outbreak detection when horizontal transmission of plasmids is the primary mode of spread, particularly due to the remarkable plasticity of plasmids, which can undergo substantial changes even within a single patient [19].

We noted significant shifts in NDM epidemiology during the study. From 2016-2018, most colonised patients had a history of travel to endemic regions such as South and Southeast Asia. Correspondingly, both the bacterial hosts and plasmids carrying NDMs were diverse with a predominance of *E. coli* and *bla*_NDM-5_, and there were no transmission events between patients detected. This contrasted with 2019-2021 when we noted increased NDM case numbers driven by patients with no travel history, which persisted despite strict travel restrictions imposed in Australia due to the COVID-19 pandemic.

Rather than being due to a single bacterial clone or plasmid, this spread was due to IncN and IncX3 epidemic plasmids. These plasmids were capable of extensive inter-strain transfer, being found in 14 and 11 bacterial strains, respectively. IncX3 plasmids have been recognised as drivers of NDM spread globally [3, 20, 21] and plasmids from our study closely matched internationally circulating plasmids. NDM IncN plasmids have previously been less recognised [22-25]. In our study, both plasmids were highly conserved across different bacterial hosts and different patients, suggesting circulation of these plasmids and acquisition by multiple patients. We also noted multiple NDM IncC plasmids, however these were more heterogenous, comprising a sub-cluster of 6 highly related plasmids across 4 patients.

Given these genomic findings, we attempted to detect healthcare-associated plasmid transmission. Despite applying liberal thresholds to increase sensitivity, we were only able to detect transmission events in 8/49 (16%) patients. The absence of spatiotemporal overlap but high genomic relatedness of plasmids may be explained by acquisition from undetected contacts/environmental reservoir within the hospital or acquisition outside of our institution (e.g. in the community or in other hospital networks). Indeed, in the three months prior, 67% of patients had a history of admission to hospital and 31% had a history of admission to the ICU. These findings raise concern about a potential widespread NDM outbreak with subsequent establishment of NDM endemicity, as noted in Tuscany in 2018-2019 [5, 26, 27].

Through our comprehensive long- and short-read sequencing of all study isolates, we were able to unravel several aspects of NDM plasmid spread. Firstly, inter-strain transfer of NDM plasmids occurred commonly and was a key driver for the increased NDM case numbers in 2019-2021. In addition, we demonstrated that inter-strain transfer likely occurred within patients, highlighting the potential for diversification of genetic settings of epidemic plasmids within patients’ microbiomes [28]. Secondly, while sophisticated plasmid clustering techniques provided much higher resolution than approaches such as *rep* typing, NDM plasmid plasticity rendered even these clusters imperfect [19]. We noted closely related plasmid backbones that carried different NDM variants (e.g. IncX3 plasmids) or NDM flanking regions, as well as mosaic plasmids that may have large structural differences but also regions that are obviously homologous to parent plasmids. Finally, in addition to conducting plasmid-level analyses, we showed that NDM spread may result from mobile genetic elements smaller than plasmids, such as the NDM transposon we identified in multiple plasmid backbones, including within the same patient. This transposon was also noted as one of the most common NDM flanking regions in a recent comprehensive analysis of global NDM plasmids [25].

These findings have important implications for use of WGS for outbreak detection where horizontal spread is the major contributor. While determining whether plasmids are likely identical is feasible (e.g. our analyses of IncN and IncX3 epidemic plasmids), defining differences between plasmids is a more complex endeavour. An objective threshold of plasmid relatedness suggestive of transmission is needed and will likely require use of multiple metrics including measures of identity (e.g. SNV analyses, average nucleotide identity), gene content and large-scale structural re-arrangements and may vary between different plasmids [29]. Circular, closed plasmid assemblies enabled by long-read sequencing will be central to these efforts [30].

There are several limitations to this work. Firstly, it was observational and based at a single centre. While we noted a diverse range of NDM plasmids and variants, these findings may not be generalisable to all NDMs. Secondly, we only conducted active surveillance in selected patients, likely limiting our detection of NDM colonisation. However, we did have systematic collection of all clinical NDM isolates with resulting accurate detection of NDM infection episodes. Finally, we used assembly approaches that result in highest-quality plasmid assemblies but plasmid assembly remains challenging and necessitates manual review and curation.

In summary, our study determined the changing dynamics of NDM spread in our institution, as the epidemiology shifted from an association with travel to likely local acquisition. Using long-read sequencing, we were able to pinpoint this to the arrival of successful epidemic NDM IncN and IncX3 plasmids that circulated concurrently and were noted in many bacterial hosts, suggesting inter-strain transfer. These epidemic plasmids remained remarkably stable across multiple bacterial strains within- and between-patients. We also noted that plasmids were capable of substantial plasticity, complicating efforts at determining plasmid transmission. These findings have important implications for the future use of long-read sequencing in detection and control of outbreaks where horizontal transmission plays a significant role.

## Data Availability

All data produced in the present study are available upon reasonable request to the authors

## Author contributions

N.M. and A.Y.P conceived the study. J.A.W., A.D., D.W.S. and A.W.J.J. designed and supervised sampling and collection of bacterial isolates. L.V.B., T.H., K.P. and G.Z.B. collected the bacterial isolates, performed bacterial characterisation and conducted whole genome sequencing of isolates. N.M. and A.D. collected clinical data. P.B. and A.J.S. conducted the outbreak investigation. N.M., J.H., B.V. and H.C. performed bioinformatics analyses. N.M. and A.Y.P. analysed all results. N.M. wrote the initial draft of the manuscript. N.M. and A.Y.P. contributed to the final version of the manuscript. All authors read and approved the manuscript.

