## Supplement for "Genomic investigation of multi-species and multi-variant blaNDM outbreak reveals key role of horizontal IncN and IncX3 plasmid transfer"

**Supplementary Methods**

*Culture, DNA extraction and sequencing*

All bacterial isolates were grown on cation-adjusted Mueller-Hinton II agar (Becton-Dickinson) at 37°C for 16 hours. Genomic DNA was extracted from bacterial plate culture using the GenFind V3 Reagent Kit (Beckman Coulter) as per manufacturer's instructions. Libraries for short read sequencing were prepared using the Nextera Flex DNA Library Prep Kit (Illumina), and 150 bp paired-end sequencing was performed on the NovaSeq 6000 system (Illumina). Libraries for long-read sequencing were prepared using the Ligation Sequencing Kit with Native Barcoding Expansion (Oxford Nanopore Technologies) and sequenced on the MinION instrument with an R9.4.1 flow cell (Oxford Nanopore Technologies) for 48 hours. Basecalling was performed with Guppy v.4.0.14 using the 'high accuracy' basecalling model.

*De novo assembly and annotation*

We constructed *de novo* assemblies incorporating short- and long-read data for all isolates. We used a long-read-first assembly approach using a bespoke pipeline (https://github.com/HughCottingham/clinopore-nf) that incorporates Flye v2.9.2 with subsequent polishing with Medaka v1.8.0, Polypolish v0.5.0 and Polca v3.4.1 [1-5]. Plasmid circularisation was assessed using Flye output. If *bla*$_{NDM}$ contigs were non-circularised, we re-assembled genomes using Unicycler v.0.4.08 with standard parameters [6]. Assembly quality was checked using Quast v5.2.0 [7] and species identification was performed using GTDB-Tk v1.0.2 [8] and checked against isolate identification performed at time of isolate collection. Genomes were annotated using Prokka v1.14.6 [9]. We then performed resistance gene and plasmid replicon detection with Abricate v.1.0.0 [10], using the NCBI Antibiotic Resistance and PlasmidFinder databases, respectively. We determined *in silico* multi-locus sequence type (ST) using 'mlst' v.2.19.0 [11]. All inconclusive ST calls with 'mlst' were checked with SRST2 v0.2.0 [12].

*Core genome-based phylogenetic analyses*

We performed core genome-based phylogenetic analyses on key STs, defined as those with ≥2 genomes available from ≥2 patients to create a core genome alignment. RefSeq genomes of the same ST were included for context in phylogenetic analyses. We chose one completed, closed assembly from our institution for each ST to use as a reference. Mobile genetic elements were excluded from these reference assemblies using PHASTER and IslandViewer 4 [13, 14]. A core chromosomal single nucleotide variant (SNV) alignment was generated using Snippy v.4.6.0 [15] and recombination was removed using Gubbins v3.3 [16]. We then used this core genome alignment in IQtree v.2.0.3 to generate maximum likelihood phylogenies for each ST [17], with the best-fit model chosen using ModelFinder [18]. For each ST, median SNV distances between isolates from our institution were then calculated.

*Plasmid analyses*

Using Abricate, we identified $bla_{\mathrm{NDM}}$-harboring contigs that were putative plasmids in our hybrid assemblies. We then used the MOB-typer v1.4.9 tool to determine plasmid replicons present, as well as to assign clusters [19]. We identified possible mosaic plasmids resulting from fusion events by examining plasmid replicon content within MOB-typer cluster and identifying plasmids which had presence of additional plasmid replicons then manually inspecting the assemblies.

We then conducted analyses within all plasmid groups within our dataset with >1 plasmid. In order to identify SNVs in the plasmid backbone, we used Snippy v.4.6.0 [15] to create a core SNV alignment by mapping short reads to a reference plasmid from our institution from each plasmid group. We used progressiveMauve v2.4.0.r4736 to align all plasmids within a plasmid group and assess for structural re-arrangements [20].

We used Flanker v0.1.5 [21] to identify and cluster flanking sequences around $bla_{\mathrm{NDM}}$. We performed clustering 5000 bp downstream of the $bla_{\mathrm{NDM}}$ gene across windows in 500 bp increments. Clustering was only performed downstream due to prior findings from Acman et al. which showed very high diversity of flanking regions upstream of $bla_{\mathrm{NDM}}$ [22]. We extracted flanking regions and performed progressiveMauve to align all flanking sequences. Geneious v10.2.6 (https://www.geneious.com) was used to visualize and assess for structural re-arrangements, with subsequent manual annotation in Adobe Illustrator v2020.24.3. We compared key plasmid groups (IncX3, IncN and IncC) from the Alfred Hospital by using a representative plasmid from each group as a reference and conducting

BLAST searches and noting the top 5 matches. For the best NDM plasmid matches, we visualized the comparisons between Alfred Hospital and publicly available plasmids in Easyfig v2.2.6 [23].

*Data availability*

Illumina/Nanopore read data were deposited in the NCBI Sequence Read Archive under project accession PRJNA967113. Completed genome assemblies were deposited in GenBank; accessions are listed in Supp. Table 1.

*Code availability*

The code generated during this study is available on GitHub (https://github.com/nenadmacesic/alfred_ndm).

| Year of admission | Days from admission to NDM | NDM colonisation status | Site of infection | Bacterial host strain | NDM variant | NDM genetic setting | Prior travel | Antimicrobial treatment regimen | Crude 7-day mortality | Crude 30-day mortality |
|---|---|---|---|---|---|---|---|---|---|---|
| 2016 | 0 | No prior NDM colonisation | Urinary tract | *Escherichia coli* ST405 | $bla_{NDM-5}$ | IncFIA (AA170 AH818) | South Asia | Amikacin/ Fosfomycin | No | No |
| 2016 | 14 | Prior NDM colonisation | Prostatitis | *Escherichia coli* ST410 | $bla_{NDM-5}$ | IncFIA (AA323 AI214) | South Asia | Tigecycline / Meropenem / Fosfomycin | No | No |
| 2018 | 8 | Prior NDM colonisation | Urinary tract | *Escherichia coli* ST405 | $bla_{NDM-5}$ | IncI-gamma/K1 | South Asia | Tigecycline / Fosfomycin | No | No |
| 2019 | 17 | Prior NDM colonisation | Pneumonia with bacteremia | *Klebsiella pneumoniae* ST15 | $bla_{NDM-4}$ | IncFII (AA450 AI539) | Southeast Asia | Ceftazidime/Avibactam / Aztreonam | No | Yes |
| 2019 | 4 | No prior NDM colonisation | Wound | *Acinetobacter baumannii* ST2 | $bla_{NDM-1}$ | Chromosome | South Asia | Trimethoprim / Sulphamethoxazole | No | No |
| 2019 | 69 | Prior NDM colonisation | Sternal wound infection | *Klebsiella variicola* ST596 | $bla_{NDM-1}$ | IncN | N/A | Ceftazidime/Avibactam/ Aztreonam / Tigecycline | No | Yes |
| 2020 | 0 | No prior NDM colonisation | Wound | *Klebsiella pneumoniae* ST16 | $bla_{NDM-1}$ | IncFIB (AA405 AI436) | Middle East | Tigecycline | No | No |
| 2020 | 13 | No prior NDM colonisation | Wound | *Pseudomonas aeruginosa* ST644 | $bla_{NDM-1}$ | Chromosome | N/A | Colistin / Meropenem | No | No |
| 2020 | 44 | Prior NDM colonisation | Pneumonia | *Klebsiella quasipneumoniae* ST5551 | $bla_{NDM-1}$ | IncN | N/A | Fosfomycin | No | No |

| | | | | | | | | | | |
|---|---|---|---|---|---|---|---|---|---|---|
| 2020 | 9 | No prior NDM colonisation | Peritonitis | *Klebsiella quasipneumoniae* ST5551 | $bla_{NDM-1}$ | IncN | N/A | Tigecycline | No | No |
| 2020 | 3 | Prior NDM colonisation | Urinary tract | *Enterobacter hormaechei* ST1015 | $bla_{NDM-1}$ | IncC | N/A | Fosfomycin / Nitrofurantoin / Ciprofloxacin | No | No |
| 2020 | 46 | No prior NDM colonisation | Wound | *Citrobacter braakii* ST567 | $bla_{NDM-7}$ | IncX3 | N/A | Trimethoprim / Sulfamethoxazole | No | No |
| 2021 | 138 | No prior NDM colonisation | Pleural fluid | *Escherichia coli* ST127 | $bla_{NDM-7}$ | IncX3 | N/A | Ciprofloxacin | No | No |
| 2021 | 17 | Prior NDM colonisation | Urinary tract | *Klebsiella pneumoniae* ST485 | $bla_{NDM-1}$ | IncN | N/A | Trimethoprim / Sulfamethoxazole | No | No |

Abbreviations: NDM – New Delhi metallo-beta-lactamase; ST – sequence type

**Supp. Table 2 - Summary of study isolates**

| Patient ID | Isolate | Species | MLST | Specimen type | Assembly type | Collection year | NDM variant | Other carbapenema-ses present | NDM genetic setting | Illumina SRA accession no. | Oxford Nanopore SRA accession no. | GenBank accession no. |
|---|---|---|---|---|---|---|---|---|---|---|---|---|
| 1 | CPO124 | *Escherichia coli* | 405 | Urine | Flye | 2016 | blaNDM-5 | | IncFIA (AA170_AH818) | SRR24474964 | SRR24475002 | JASUUY000000000 |
| 2 | CPO125 | *Escherichia coli* | 410 | Rectal swab | Flye | 2016 | blaNDM-5 | | IncFIA (AA323_AI214) | SRR24474921 | SRR24475001 | JASUUZ000000000 |
| 3 | CPO126 | *Klebsiella pneumoniae* | 14 | Rectal swab | Flye | 2016 | blaNDM-1 | | IncFIB (AA018_AH560) | SRR24474910 | SRR24475000 | JASUVA000000000 |
| 3 | CPO122 | *Escherichia coli* | 410 | Urine | Flye | 2016 | blaNDM-5 | blaOXA-181 | IncFIA (AA323_AI214) | SRR24474975 | SRR24475003 | JASUUX000000000 |
| 4 | CPO114 | *Escherichia coli* | 394 | Urine | Flye | 2017 | blaNDM-1 | | IncFIA (AA327_AI227) \| Chromosome | SRR24474986 | SRR24475004 | JASUUW000000000 |
| 5 | CPO099 | *Escherichia coli* | 405 | Rectal swab | Flye | 2017 | blaNDM-5 | | IncFIA (AA324_AI219) | SRR24475025 | SRR24475005 | JASUUV000000000 |
| 6 | CPO097 | *Escherichia coli* | 648 | Rectal swab | Unicycler | 2017 | blaNDM-7 | | IncX3 (AA038_AH615) | SRR24475006 | SRR24475007 | JASUWW000000000 |
| 6 | CPO096 | *Escherichia coli* | 46 | Rectal swab | Flye | 2017 | blaNDM-5 | | IncFIA (AA172_AH832) | SRR24475017 | SRR24475008 | JASUUU000000000 |
| 7 | CPO090 | *Escherichia coli* | 410 | Rectal swab | Flye | 2018 | blaNDM-5 | | IncFIA (AA323_AI214) | SRR24474872 | SRR24475009 | JASUUT000000000 |
| 8 | CPO084 | *Escherichia coli* | 405 | Rectal swab | Flye | 2018 | blaNDM-5 | | IncFIA (AA170_AH818) | SRR24474883 | SRR24475010 | JASUUS000000000 |
| 8 | CPO083 | *Escherichia coli* | 405 | Urine | Flye | 2018 | blaNDM-5 | | IncFIA (AA170_AH818) | SRR24474894 | SRR24475011 | JASUUR000000000 |
| 8 | CPO080 | *Escherichia coli* | 405 | Rectal swab | Flye | 2018 | blaNDM-5 | | IncI-gamma/K1 (AC026_AL043) | SRR24474937 | SRR24475012 | JASUUQ000000000 |

| | | | | | | | | | | | | |
|---|---|---|---|---|---|---|---|---|---|---|---|---|
| 9 | CPO077 | *Escherichia coli* | 405 | Urine | Flye | 2018 | blaNDM-5 | | IncI-gamma/K1 (AC026_AL043) | SRR24474948 | SRR24475013 | JASUUP000000000 |
| 10 | CPO070 | *Escherichia coli* | 1284 | Rectal swab | Flye | 2018 | blaNDM-5 | | IncX3 (AA038_AH615) | SRR24474959 | SRR24475014 | JASUUO000000000 |
| 11 | CPO053 | *Enterobacter hormaechei* | 114 | Rectal swab | Unicycler | 2018 | blaNDM-7 | blaIMP-4 | IncX3 (AA038_AH615) | SRR23384660 | SRR23384664 | GCA_030187275.1 |
| 11 | CPO035 | *Escherichia coli* | 8346 | Rectal swab | Flye | 2019 | blaNDM-5 | | IncFIA (AA170_AH820) | SRR24475027 | SRR24475016 | JASUUM000000000 |
| 12 | CPO036 | *Klebsiella pneumoniae* | 16 | Rectal swab | Flye | 2019 | blaNDM-5 | | IncX3 (AA038_AH615) | SRR24475026 | SRR24475015 | JASUUN000000000 |
| 13 | CPO195 | *Escherichia coli* | 361 | Urine | Unicycler | 2019 | blaNDM-5 | | IncX3 (AA038_AH615) | SRR24474963 | SRR24474999 | JASUWX000000000 |
| 13 | CPO206 | *Klebsiella pneumoniae* | 656 | Rectal swab | Flye | 2019 | blaNDM-4 | | IncFII (AA450_AI539) | SRR24474962 | SRR24474998 | JASUVB000000000 |
| 14 | CPO217 | *Klebsiella pneumoniae* | 15 | Urine | Flye | 2019 | blaNDM-4 | | IncFII (AA450_AI539) | SRR24474961 | SRR24474997 | JASUVC000000000 |
| 14 | CPO218 | *Klebsiella pneumoniae* | 15 | Blood Peripheral | Flye | 2019 | blaNDM-4 | | IncFII (AA450_AI539) | SRR24474960 | SRR24474996 | JASUVD000000000 |
| 15 | CPO219 | *Escherichia coli* | 405 | Rectal swab | Flye | 2019 | blaNDM-5 | | IncFIA (AA170_AH818) | SRR24474958 | SRR24475024 | JASUVE000000000 |
| 16 | CPO225 | *Enterobacter hormaechei* | 114 | Rectal swab | Unicycler | 2019 | blaNDM-7 | | IncX3 (AA038_AH615) | SRR24474957 | SRR24474995 | JASUWY000000000 |
| 17 | CPO228 | *Acinetobacter baumannii* | 2 | Wound | Flye | 2019 | blaNDM-1 | blaOXA-23 \| blaOXA-66 | Chromosome | SRR24474956 | SRR24474994 | JASUVF000000000 |
| 18 | CPO230 | *Escherichia coli* | 90 | Rectal swab | Flye | 2019 | blaNDM-5 | | IncFIA (AA170_AH818) | SRR24474955 | SRR24474993 | JASUVG000000000 |
| 19 | CPO236 | *Escherichia coli* | 176 | Rectal swab | Flye | 2019 | blaNDM-1 | | IncC (AA860_AJ272) | SRR24474954 | SRR24474992 | JASUVH000000000 |
| 20 | CPO266 | *Klebsiella michiganensis* | Novel | Rectal swab | Unicycler | 2019 | blaNDM-1 | | IncN (AA552_AI757) | SRR24474953 | SRR24474991 | JASUWZ000000000 |

| | | | | | | | | | | | | |
|---|---|---|---|---|---|---|---|---|---|---|---|---|
| 21 | CPO282 | *Klebsiella michiganensis* | Novel | Rectal swab | Flye | 2019 | blaNDM-1 | | IncN (AA552_AI757) | SRR24474952 | SRR24474990 | JASUVI000000000 |
| 22 | CPO293 | *Klebsiella variicola* | 596 | Wound | Unicycler | 2019 | blaNDM-1 | | IncN (AA552_AI757) | SRR24474951 | SRR24474989 | JASUXA000000000 |
| 23 | CPO299 | *Enterobacter hormaechei* | 88 | Rectal swab | Flye | 2019 | blaNDM-1 | | IncN (AA552_AI757) | SRR24474950 | SRR24474988 | JASUVJ000000000 |
| 24 | CPO308 | *Escherichia coli* | 648 | Rectal swab | Unicycler | 2020 | blaNDM-5 | | IncX3 (AA038_AH615) | SRR24474949 | SRR24474987 | JASUXB000000000 |
| 24 | CPO310 | *Klebsiella quasipneumoniae* | 5551 | Rectal swab | Flye | 2020 | blaNDM-1 | | IncN (AA552_AI757) | SRR24474947 | SRR24474985 | JASUVK000000000 |
| 24 | CPO311 | *Citrobacter amaloticus* | - | Rectal swab | Flye | 2020 | blaNDM-1 | | IncN (AA552_AI757) | SRR24474946 | SRR24474984 | JASUVL000000000 |
| 25 | CPO315 | *Klebsiella pneumoniae* | 16 | Wound | Flye | 2020 | blaNDM-1 | | IncFIB (AA405_AI436) | SRR24474945 | SRR24474983 | JASUVM000000000 |
| 26 | CPO316 | *Enterobacter cloacae* | 610 | Rectal swab | Flye | 2020 | blaNDM-1 | | IncN (AA552_AI757) | SRR24474944 | SRR24474982 | JASUVN000000000 |
| 27 | CPO318 | *Klebsiella pneumoniae* | 16 | Rectal swab | Unicycler | 2020 | blaNDM-1 | | IncFIB (AA405_AI436) | SRR24474943 | SRR24474981 | JASUXC000000000 |
| 28 | CPO319 | *Klebsiella pneumoniae* | 16 | Urine | Unicycler | 2020 | blaNDM-1 | | IncFIB (AA405_AI436) | SRR24474942 | SRR24474980 | JASUXD000000000 |
| 28 | CPO383 | *Pseudomos aeruginosa* | 644 | Burn | Flye | 2020 | blaNDM-1 | blaIMP-62 | Chromosome | SRR24474892 | SRR24474930 | JASUWB000000000 |
| 29 | CPO326 | *Klebsiella quasipneumoniae* | 5551 | Sputum | Flye | 2020 | blaNDM-1 | | IncN (AA552_AI757) | SRR24474941 | SRR24474979 | JASUVO000000000 |
| 29 | CPO384 | *Pseudomos aeruginosa* | 644 | Faeces | Flye | 2020 | blaNDM-1 | blaIMP-62 | Chromosome | SRR24474891 | SRR24474929 | JASUWC000000000 |
| 29 | CPO430 | *Pseudomos aeruginosa* | 644 | Faeces | Flye | 2020 | blaNDM-1 | blaIMP-62 | Chromosome | SRR24474882 | SRR24474920 | JASUWJ000000000 |
| 29 | CPO443 | *Pseudomos aeruginosa* | 644 | Faeces | Flye | 2020 | blaNDM-1 | | Chromosome | SRR24474874 | SRR24474912 | JASUWM000000000 |
| 29 | CPO327 | *Klebsiella pneumoniae* | 11 | Wound | Flye | 2020 | blaNDM-5 | blaOXA-48 | IncFIB (AA405_AI436) | SRR24474940 | SRR24474978 | JASUVP000000000 |
| 29 | CPO329 | *Citrobacter amaloticus* | - | Faeces | Flye | 2020 | blaNDM-1 | | IncN (AA552_AI757) | SRR24474939 | SRR24474977 | JASUVQ000000000 |
| 29 | CPO330 | *Klebsiella quasipneumoniae* | 5551 | Faeces | Flye | 2020 | blaNDM-1 | | IncN (AA552_AI757) | SRR24474938 | SRR24474976 | JASUVR000000000 |
| 30 | CPO331 | *Klebsiella quasipneumoniae* | 5551 | Rectal swab | Flye | 2020 | blaNDM-1 | | IncN (AA552_AI757) | SRR24474936 | SRR24474974 | JASUVS000000000 |

| | | | | | | | | | | | | |
|---|---|---|---|---|---|---|---|---|---|---|---|---|
| 31 | CPO340 | *Klebsiella quasipneumoniae* | 5551 | Rectal swab | Flye | 2020 | blaNDM-1 | | IncN (AA552_AI757) | SRR24474933 | SRR24474971 | JASUVU000000000 |
| 31 | CPO334 | *Klebsiella quasipneumoniae* | 5551 | Urine | Flye | 2020 | blaNDM-1 | | IncN (AA552_AI757) | SRR24474935 | SRR24474973 | JASUVT000000000 |
| 32 | CPO346 | *Raoultella ornithinolytica* | - | Rectal swab | Flye | 2020 | blaNDM-1 | | IncN (AA552_AI757) | SRR24474932 | SRR24474970 | JASUVV000000000 |
| 33 | CPO336 | *Escherichia coli* | 131 | Rectal swab | Unicycler | 2020 | blaNDM-1 | | IncN (AA552_AI757) | SRR24474934 | SRR24474972 | JASUXE000000000 |
| 33 | CPO350 | *Citrobacter freundii* | 64 | Rectal swab | Flye | 2020 | blaNDM-1 | | IncN (AA552_AI757) | SRR24474899 | SRR24474969 | JASUVW000000000 |
| 33 | CPO351 | *Enterobacter hormaechei* | 1015 | Rectal swab | Flye | 2020 | blaNDM-1 | | IncC (AA860_AJ272) | SRR24474898 | SRR24474968 | JASUVX000000000 |
| 34 | CPO352 | *Klebsiella pneumoniae* | 16 | Swab | Flye | 2020 | blaNDM-1 | | IncFIB (AA405_AI436) | SRR24474897 | SRR24474967 | JASUVY000000000 |
| 35 | CPO354 | *Klebsiella pneumoniae* | 17 | Rectal swab | Unicycler | 2020 | blaNDM-7 | | IncX3 (AA038_AH615) | SRR24474896 | SRR24474966 | JASUXF000000000 |
| 36 | CPO406 | *Citrobacter braakii* | Novel | Rectal swab | Unicycler | 2020 | blaNDM-7 | | IncX3 (AA038_AH615) | SRR24474886 | SRR24474924 | JASUXG000000000 |
| 36 | CPO363 | *Klebsiella quasipneumoniae* | 5551 | Peritoneal fluid | Flye | 2020 | blaNDM-1 | | IncN (AA552_AI757) | SRR24474893 | SRR24474931 | JASUWA000000000 |
| 36 | CPO358 | *Klebsiella pneumoniae* | 16 | Urine | Flye | 2020 | blaNDM-1 | | IncFIB (AA405_AI436) | SRR24474895 | SRR24474965 | JASUVZ000000000 |
| 37 | CPO371 | *Citrobacter freundii* | 492 | Rectal swab | Unicycler | 2020 | blaNDM-7 | blaIMP-4 | IncX3 (AA038_AH615) | SRR23384825 | SRR23384576 | GCA_030184215.1 |
| 37 | CPO408 | *Klebsiella pneumoniae* | 17 | Rectal swab | Flye | 2020 | blaNDM-7 | blaIMP-4 | IncX3 (AA038_AH615) | SRR24474885 | SRR24474923 | JASUWH000000000 |
| 37 | CPO387 | *Escherichia coli* | 940_1LV | Swab | Flye | 2020 | blaNDM-5 | | IncX3 (AA038_AH615) | SRR24474890 | SRR24474928 | JASUWD000000000 |
| 38 | CPO390 | *Enterobacter hormaechei* | 1015 | Rectal swab | Flye | 2020 | blaNDM-1 | | IncC (AA860_AJ272) | SRR24474889 | SRR24474927 | JASUWE000000000 |
| 38 | CPO391 | *Enterobacter hormaechei* | 1015 | Urine | Flye | 2020 | blaNDM-1 | | IncC (AA860_AJ272) | SRR24474888 | SRR24474926 | JASUWF000000000 |

| | | | | | | | | | | | | |
|---|---|---|---|---|---|---|---|---|---|---|---|---|
| 39 | CPO404 | *Klebsiella michiganensis* | 85_2LV | Rectal swab | Flye | 2020 | blaNDM-1 | | IncN (AA552_AI757) | SRR24474887 | SRR24474925 | JASUWG000000000 |
| 40 | CPO428 | *Citrobacter braakii* | 567 | Tissue | Flye | 2020 | blaNDM-7 | | IncX3 (AA038_AH615) | SRR24474884 | SRR24474922 | JASUWI000000000 |
| 40 | CPO429 | *Klebsiella aerogenes* | 304 | Urine | Flye | 2020 | blaNDM-1 | blaIMP-4 | IncN (AA552_AI757) | SRR23384815 | SRR23384532 | GCA_030183875.1 |
| 40 | CPO433 | *Klebsiella michiganensis* | 85_2LV | Rectal swab | Unicycler | 2020 | blaNDM-1 | | IncN (AA552_AI757) | SRR24474881 | SRR24474919 | JASUXH000000000 |
| 40 | CPO434 | *Klebsiella pneumoniae* | 17 | Urine | Flye | 2020 | blaNDM-7 | | IncX3 (AA038_AH615) | SRR24474880 | SRR24474918 | JASUWK000000000 |
| 40 | CPO436 | *Enterobacter hormaechei* | 1015 | Rectal swab | Flye | 2020 | blaNDM-1 | | IncC (AA860_AJ272) | SRR24474879 | SRR24474917 | JAUBKK000000000 |
| 41 | CPO438 | *Escherichia coli* | 967 | Swab | Unicycler | 2020 | blaNDM-1 | | IncC (AA860_AJ272) | SRR24474878 | SRR24474916 | JASUXI000000000 |
| 41 | CPO440 | *Escherichia coli* | 1193 | Rectal swab | Unicycler | 2021 | blaNDM-1 | | IncC (AA860_AJ272) | SRR24474877 | SRR24474915 | JASUXJ000000000 |
| 41 | CPO441 | *Escherichia coli* | 1193 | Rectal swab | Unicycler | 2021 | blaNDM-1 | | IncC (AA860_AJ272) | SRR24474876 | SRR24474914 | JASUXK000000000 |
| 41 | CPO442 | *Klebsiella pneumoniae* | 16 | Rectal swab | Flye | 2021 | blaNDM-1 | blaOXA-232 | IncFIA (AA170_AH820) | SRR24474875 | SRR24474913 | JASUWL000000000 |
| 42 | CPO446 | *Escherichia coli* | 127 | Pleural fluid | Unicycler | 2021 | blaNDM-7 | | IncX3 (AA038_AH615) | SRR24474873 | SRR24474911 | JASUXL000000000 |
| 43 | CPO449 | *Escherichia coli* | 127 | BAL | Unicycler | 2021 | blaNDM-7 | | IncX3 (AA038_AH615) | SRR24474869 | SRR24474907 | JASUXM000000000 |
| 44 | CPO447 | *Enterobacter hormaechei* | 1015 | Swab | Flye | 2021 | blaNDM-1 | | IncC (AA860_AJ272) | SRR24474871 | SRR24474909 | JASUWN000000000 |
| 44 | CPO448 | *Enterobacter hormaechei* | 1015 | Rectal swab | Flye | 2021 | blaNDM-1 | | IncC (AA860_AJ272) | SRR24474870 | SRR24474908 | JASUWO000000000 |

| | | | | | | | | | | | |
|---|---|---|---|---|---|---|---|---|---|---|---|
| 45 | CPO450 | *Citrobacter braakii* | 567 | Rectal swab | Flye | 2021 | blaNDM-7 | IncX3 (AA038_AH615) | SRR24474868 | SRR24474906 | JASUWP000000000 |
| 46 | CPO452 | *Citrobacter youngae* | 187 | Rectal swab | Flye | 2021 | blaNDM-1 | IncN (AA552_AI757) | SRR24475023 | SRR24474905 | JASUWQ000000000 |
| 47 | CPO453 | *Klebsiella michiganensis* | Novel | Rectal swab | Flye | 2021 | blaNDM-1 | IncN (AA552_AI757) | SRR24475022 | SRR24474904 | JASUWR000000000 |
| 48 | CPO591 | *Klebsiella pneumoniae* | 485 | Rectal swab | Flye | 2021 | blaNDM-1 | IncN (AA552_AI757) | SRR24475021 | SRR24474903 | JASUWS000000000 |
| 48 | CPO593 | *Escherichia coli* | 1284 | Rectal swab | Flye | 2021 | blaNDM-5 | IncFIA (AA324_AI219) | SRR24475019 | SRR24474901 | JASUWU000000000 |
| 48 | CPO592 | *Klebsiella pneumoniae* | 485 | Urine | Flye | 2021 | blaNDM-1 | IncN (AA552_AI757) | SRR24475020 | SRR24474902 | JASUWT000000000 |
| 49 | CPO595 | *Klebsiella michiganensis* | 29 | Urine | Flye | 2021 | blaNDM-1 | IncN (AA552_AI757) | SRR24475018 | SRR24474900 | JASUWV000000000 |

Abbreviations: ID - identification, LV - locus variant, MLST - Multi-locus sequence type, NDM - New Delhi metallo-beta-lactamase, SRA - Sequence Read Archive.

**Supp. Table 3 – Pairwise SNV distances of key New Delhi metallo-beta-lactamase bacterial host strains**

| Bacterial strain | Total genomes | Median pairwise SNV distance | Interquartile range | Total patients | Minimum between-patient pairwise SNV distance |
|---|---|---|---|---|---|
| *Escherichia coli* ST405 | 7 | 121 | 103 – 300 | 5 | 32 |
| *Klebsiella pneumoniae* ST16 | 7 | 134.5 | 4 – 171.25 | 4 | 0 |
| *Klebsiella quasipneumoniae* ST5551 | 7 | 8 | 4 – 11 | 3 | 2 |
| *Enterobacter hormaechei* ST1015 | 6 | 4 | 3 – 5 | 4 | 3 |
| *Escherichia coli* ST410 | 3 | 78 | 28.75 – 110 | 2 | 110 |
| *Klebsiella michiganensis* - Novel ST | 3 | 24 | 12 – 24 | 2 | 24 |
| *Klebsiella pneumoniae* ST17 | 3 | 2 | 1.5 – 2.5 | 2 | 2 |
| *Citrobacter braakii* ST567 | 2 | 1* | | 2 | 1 |
| *Enterobacter hormaechei* ST114 | 2 | 1847* | | 2 | 1847 |
| *Escherichia coli* ST1284 | 2 | 3417* | | 2 | 3417 |
| *Escherichia coli* ST648 | 2 | 11845* | | 2 | 11845 |
| *Klebsiella michiganensis* ST85 (2LV) | 2 | 4* | | 2 | 4 |

*Pairwise distance reported as only two genomes available for analyses

Abbreviations: LV – locus variant; SNV – single nucleotide variant; ST – sequence type

**Supp. Table 4 – Pairwise SNV distances of key New Delhi metallo-beta-lactamase plasmid groups**

| Plasmid group (MOBtyper cluster) | Median pairwise SNV distance | Mean pairwise SNV distance | Interquartile range |
|---|---|---|---|
| IncC (AA860 AJ272) | 1 | 8.2 | 0 - 22 |
| IncFIA (AA170 AH818) | 835 | 768.5 | 647.25 - 1115 |
| IncFIA (AA170 AH820) | 196* | | |
| IncFIA (AA323 AI214) | 228 | 152.6 | 114.5 - 228.5 |
| IncFIA (AA324 AI219) | 41* | | |
| IncFIB (AA405 AI436) | 0 | 47.7 | 0 - 143 |
| IncFII (AA450 AI539) | 267 | 178 | 133.5 - 267 |
| IncI-gamma/K1 (AC026 AL043) | 4* | | |
| IncN (AA552 AI757) | 0 | 0.08 | 0 - 0 |
| IncX3 (AA038 AH615) | 2 | 2.2 | 0 - 3 |

*Pairwise distance reported as only two genomes available for analyses

Abbreviations: SNV – single nucleotide variant

**Supp. Table 5 – BLAST matches for Alfred Hospital epidemic plasmids**

| Reference plasmid | Description | Max Score | Query Coverage | E value | Percentage identity | GenBank accession |
|---|---|---|---|---|---|---|
| Alfred Hospital IncX3 | *Escherichia coli* strain EC-YC1908-399 plasmid p399-4, complete sequence | 85148 | 100% | 0 | 99.96 | CP084538.1 |
| | *Escherichia coli* EC08 plasmid pEC08_NDM5 DNA, complete sequence | 84987 | 100% | 0 | 99.9 | LC521848.1 |
| | *Escherichia coli* strain CFSAN064035 plasmid pGMI17-003_4, complete sequence | 84108 | 100% | 0 | 99.94 | CP031138.1 |
| | *Klebsiella quasipneumoniae* strain JNQH473 plasmid pJNQH473-4, complete sequence | 84051 | 100% | 0 | 99.95 | CP075887.1 |
| | *Klebsiella pneumoniae* strain KpN01 plasmid pKpN01-NDM7, complete sequence | 83534 | 100% | 0 | 99.95 | CP012990.1 |
| Alfred Hospital IncN | *Citrobacter freundii* strain Cf7308 plasmid pNDM-Cf7308, complete sequence | 53033 | 95% | 0 | 99.93 | CP092465.1 |
| | *Enterobacter asburiae* strain 5549 plasmid IncN_1, complete sequence | 53033 | 95% | 0 | 99.93 | CP093156.1 |
| | *Escherichia coli* strain Z30 plasmid pZ30-NDM-29, complete sequence | 53033 | 95% | 0 | 99.93 | CP066846.1 |
| | *Enterobacter cloacae* strain CBG15936 plasmid pNDM1-CBG, complete sequence | 53029 | 95% | 0 | 99.93 | CP046118.1 |
| | *Klebsiella pneumoniae* strain FF1009 plasmid pFF1009_1, complete sequence | 52951 | 91% | 0 | 99.95 | CP077825.1 |
| Alfred Hospital IncC | *Klebsiella pneumoniae* isolate BB1465 genome assembly, plasmid: pKP-CTX-M-15_ | 1.77E+05 | 92% | 0 | 99.95 | LR822059.1 |

| | | | | | | |
|---|---|---|---|---|---|---|
| | *Proteus mirabilis* strain AR_0159 plasmid tig00000137, complete sequence | 1.76E+05 | 93% | 0 | 99.99 | CP021551.1 |
| | *Citrobacter werkmanii* isolate BB1459 genome assembly, plasmid: pCW-CTX-M-15B_ | 1.75E+05 | 94% | 0 | 99.97 | LR822057.1 |
| | *Escherichia coli* strain Ecol_732 plasmid pEC732_IMP14, complete sequence | 1.70E+05 | 89% | 0 | 99.95 | CP015139.1 |
| | *Klebsiella michiganensis* strain BD-50-Km plasmid pBD-50-Km_VIM-1, complete sequence | 1.60E+05 | 94% | 0 | 99.98 | CP061931.1 |

**Supp. Table 6 – Within-patient plasmid pairwise single nucleotide variant distances**

| Plasmid group (MOBtyper cluster) | Total within-patient pairs | Median SNV distance | Mean SNV distance | Interquartile range | Range |
|---|---|---|---|---|---|
| *bla*$_{NDM-1}$ IncC (AA860 AJ272) | 3 | 0 | 0 | 0 - 0 | 0 - 0 |
| *bla*$_{NDM-5}$ IncFIA (AA323 AI214) | 1 | 1* | | | |
| *bla*$_{NDM-1}$ IncFIB (AA405 AI436) | 10 | 0 | 57.2 | 0 – 143 | 0 - 143 |
| *bla*$_{NDM-4}$ IncFII (AA450 AI539) | 3 | 267 | 178 | 133.5 - 267 | 0 - 267 |
| *bla*$_{NDM-5}$ IncI-gamma/K1 (AC026 AL043) | 1 | 4* | | | |
| *bla*$_{NDM-1}$ IncN (AA552 AI757) | 27 | 0 | 0 | 0 - 0 | 0 - 0 |
| *bla*$_{NDM-7}$ IncX3 (AA038 AH615) | 7 | 0 | 0 | 0 - 0 | 0 - 0 |

*Pairwise distance reported as only two genomes available for analyses

Abbreviations: SNV – single nucleotide variant

**Supp. Figure 1- Epidemiological curves of *bla*NDM 2016-2021**

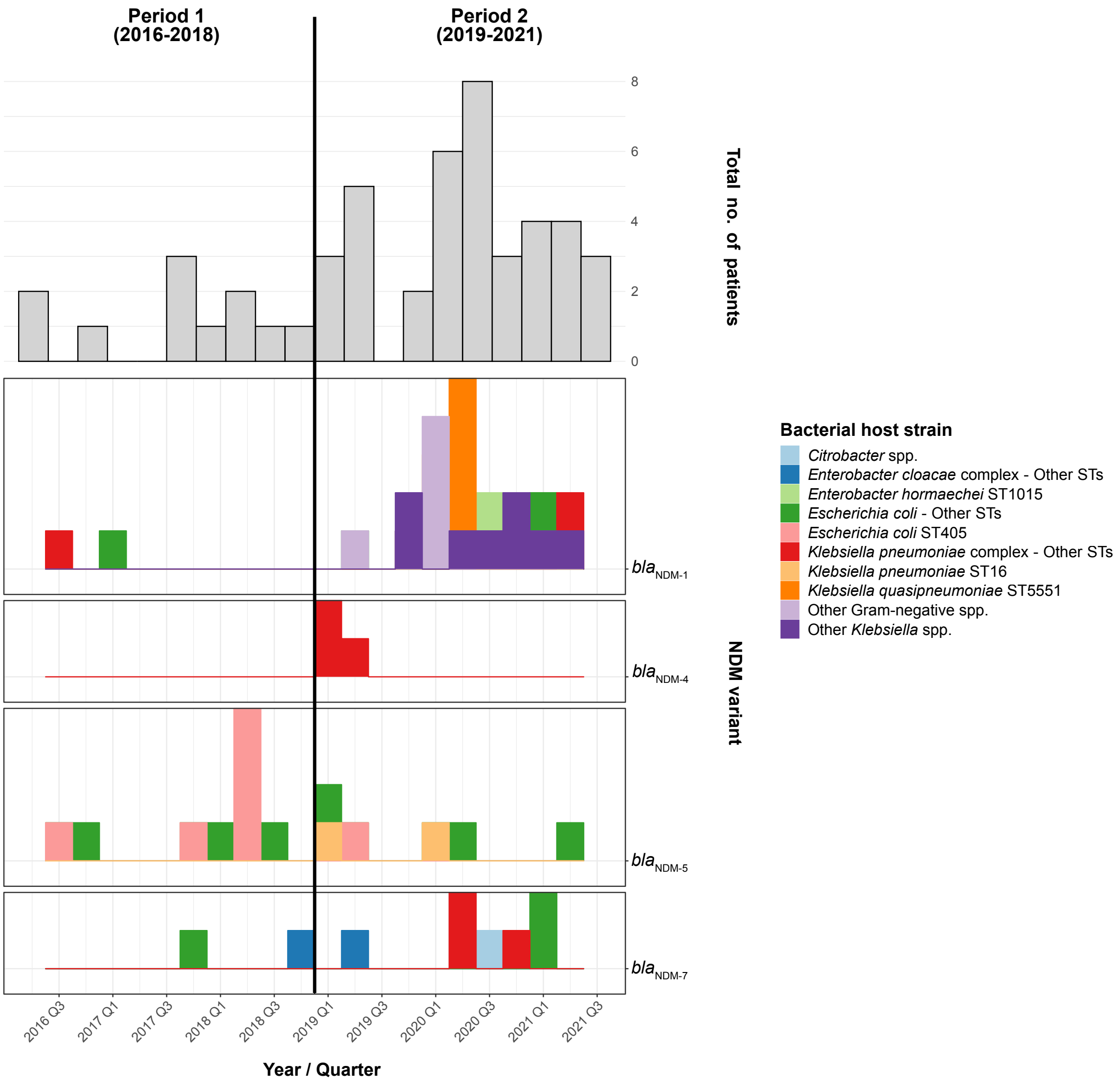


Top panel indicates overall number of NDM patients during each year/quarter. Bottom panels show *bla*NDM genomes per NDM variant (shown as separate panels) and bacterial host strain (shown as different colours).

Abbreviations: NDM – New Delhi metallo-beta-lactamase; No. – number; ST – sequence type.

**Supp. Figure 2 – Comparison of Alfred Hospital and global New Delhi metallo-beta-lactamase plasmids**

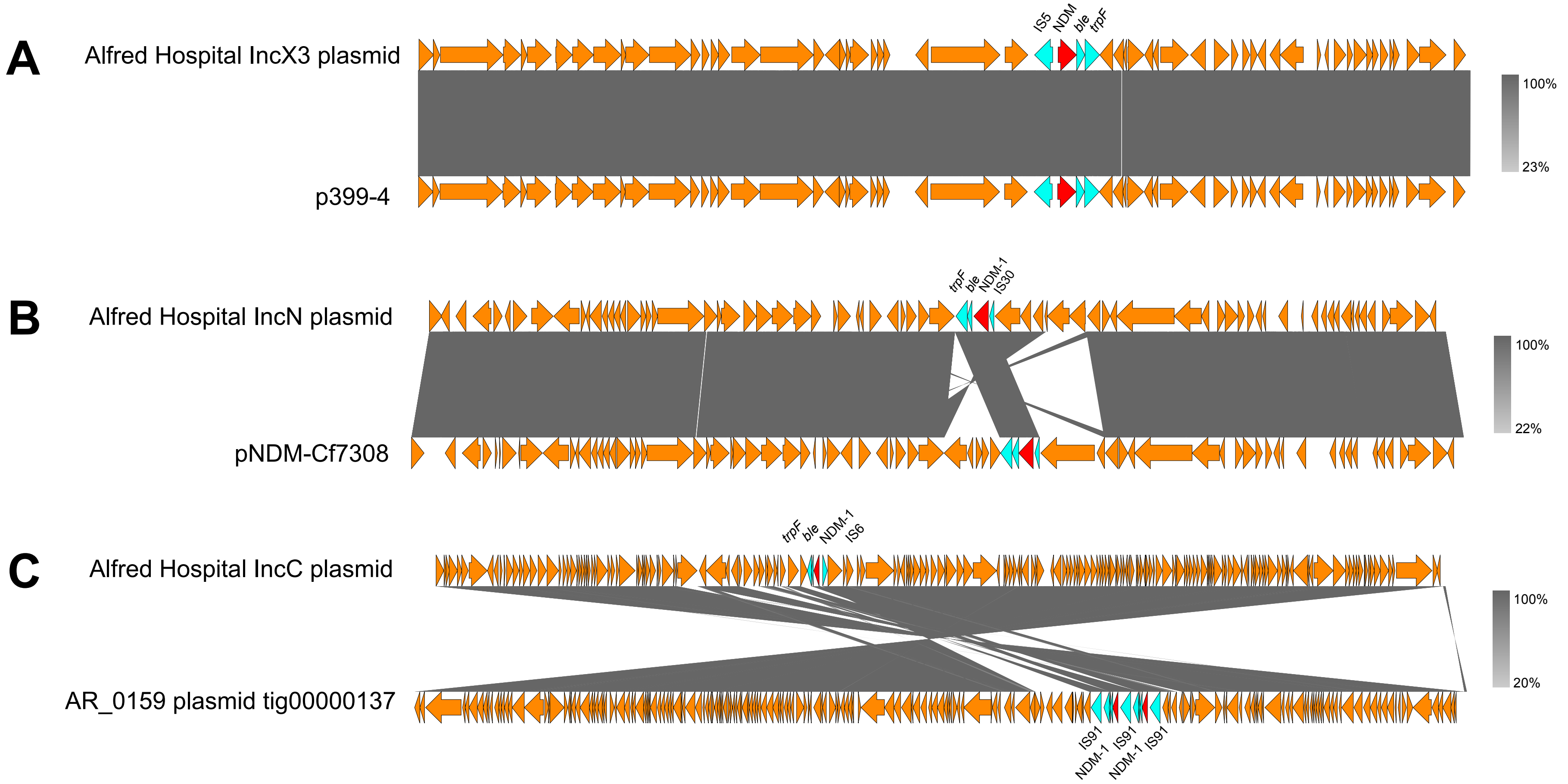


We conducted comparative analyses with global New Delhi metallo-beta-lactamase (NDM) plasmids that were closely related to NDM epidemic plasmids from our study. The location of $bla_{NDM}$ genes is shown in red, with the genetic context in light blue. Panel A shows that IncX3 plasmids from our study were almost identical to global NDM IncX3 plasmids (100% coverage and 99.96% identity). Panels B and C demonstrate IncN and IncC plasmids, respectively. Plasmids from our study had more significant differences to NDM global plasmids, with IncN plasmids having 95% coverage and 99.93% identity while IncC plasmids had 93% coverage and 99.99% identity.

**Supp. Figure 3 – Comparative analysis of non-epidemic NDM plasmids**

**IncFIA-type plasmids (AA170 AH818)**

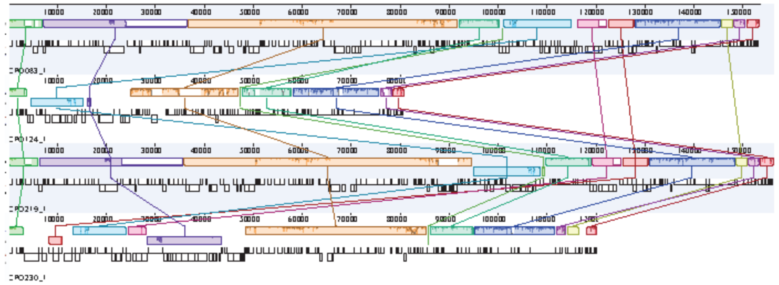

**IncFIA-type plasmids (AA324 AI219)**

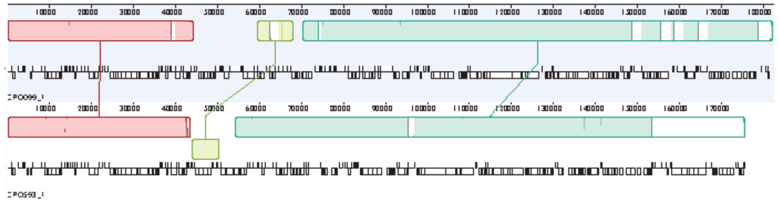

**IncFIA-type plasmids (AA170 AH820)**

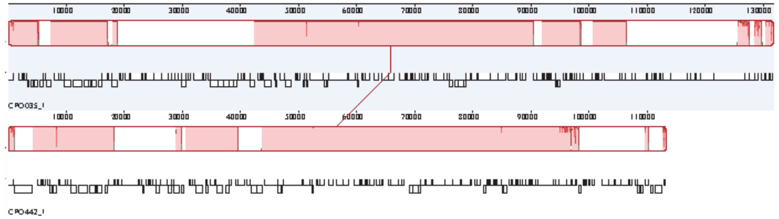

**IncFIB-type plasmids (AA405 AI436)**

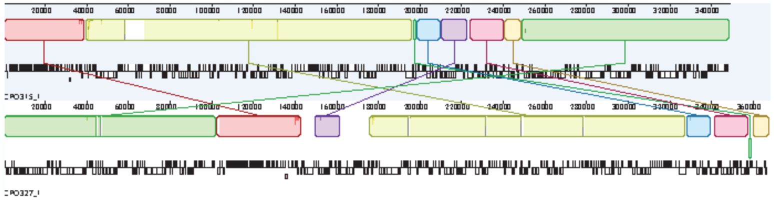

**IncFIA-type plasmids (AA323 AI214)**

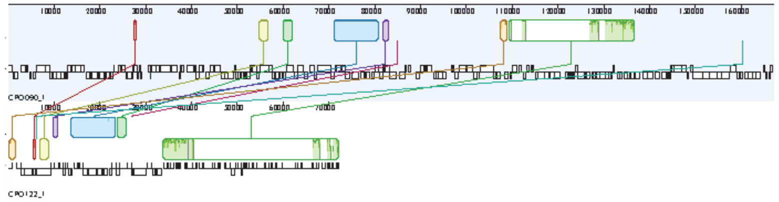

We conducted comparative analysis of non-epidemic NDM plasmid groups from our study from which multiple plasmids were available for analysis. Each colored field represents a locally collinear block, a homologous region of sequence shared by multiple plasmids without any rearrangement of that region. Same colours indicate the same regions present in different plasmids. In contast to epidemic plasmids, NDM non-epidemic plasmids showed more large-scale structural differences despite plasmids being classified within the same plasmid groups.

Abbreviations: NDM – New Delhi metallo-beta-lactamase; ST – sequence type.

**Supp. Figure 4 – Promiscuous NDM transposon found in seven plasmid groups**

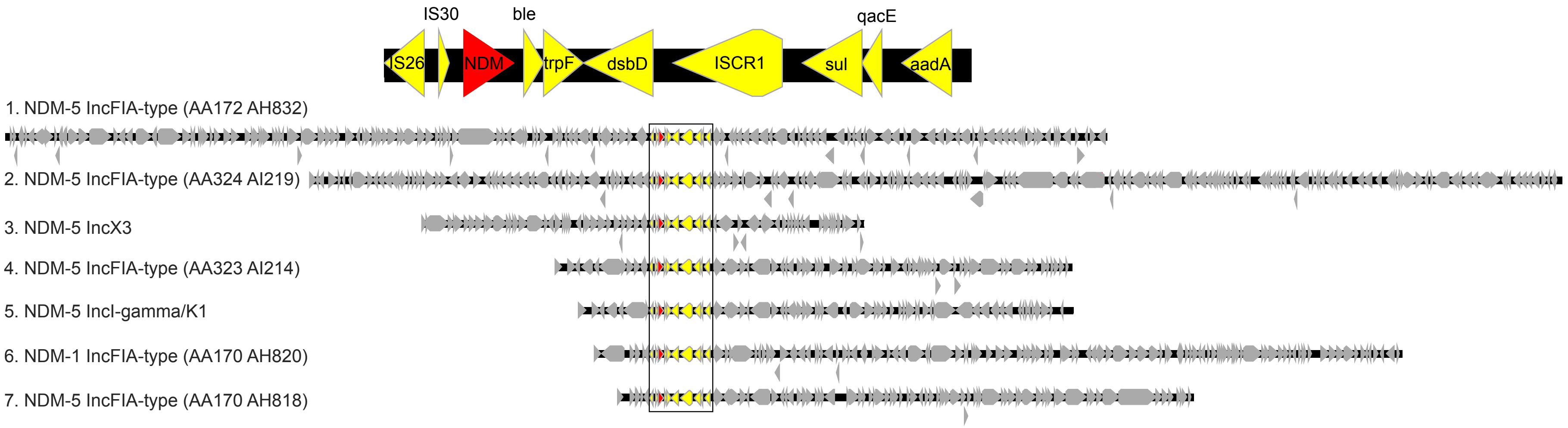


The top of the figure shows the genes contained within the transposon, including *bla*NDM. The bottom of the figure shows the transposon (in colour) inserting into diverse plasmids from seven different plasmid groups. This was suggestive of movement of the transposon between different plasmid backbones.

Abbreviations: NDM – New Delhi metallo-beta-lactamase

**Supp. Figure 5 – Patients colonised with multiple NDM plasmids**

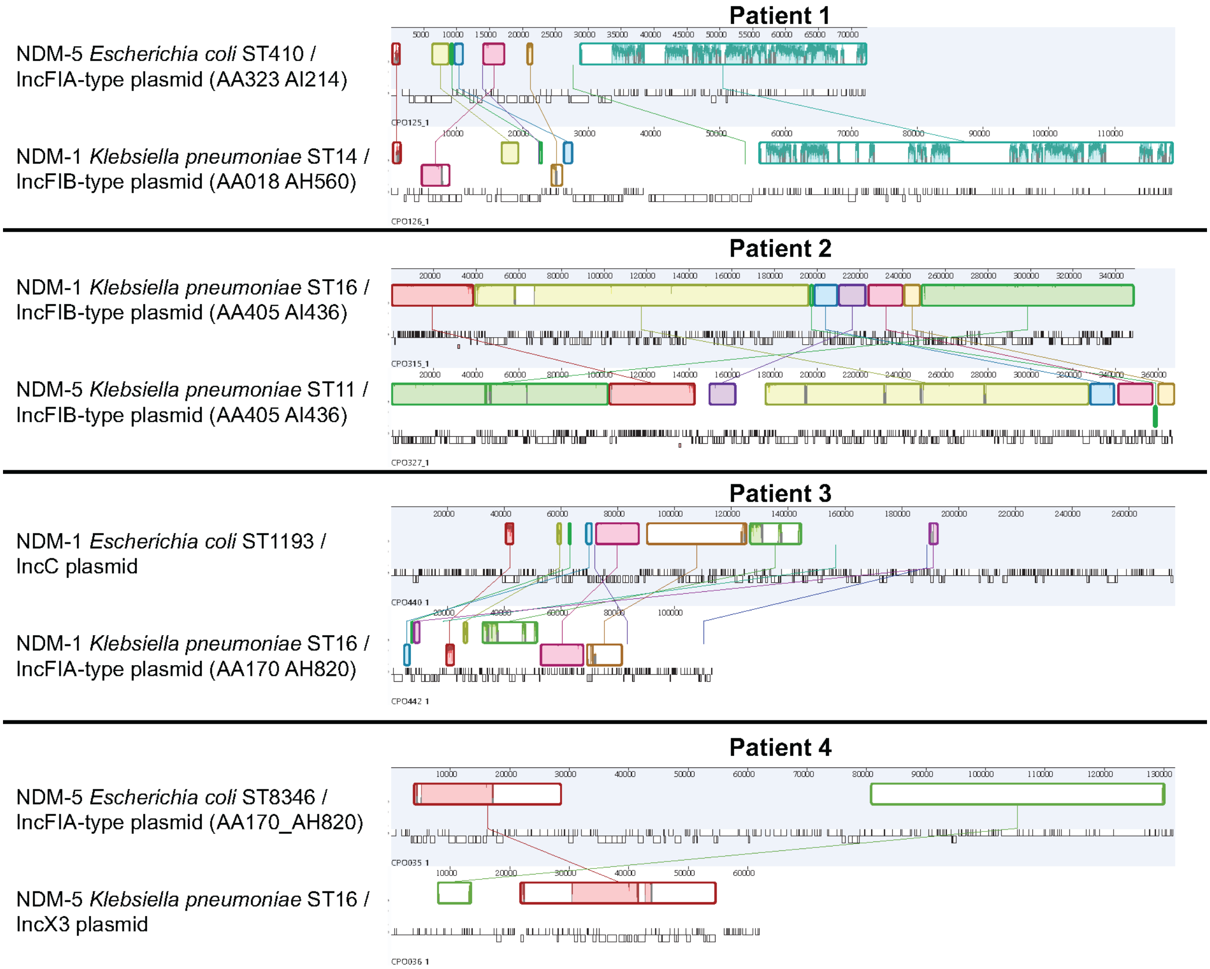


We analyzed individual patients colonised with multiple NDM plasmids as evidenced by colonisation with different NDM plasmid groups or different NDM variants. Each colored field represents a locally collinear block, a homologous region of sequence shared by multiple plasmids without any rearrangement of that region. Same colours indicate the same regions present in different plasmids.

Abbreviations: NDM – New Delhi metallo-beta-lactamase; ST – Sequence type.

**Supp. Figure 6 – Analysis of potential NDM transmission events between patients**

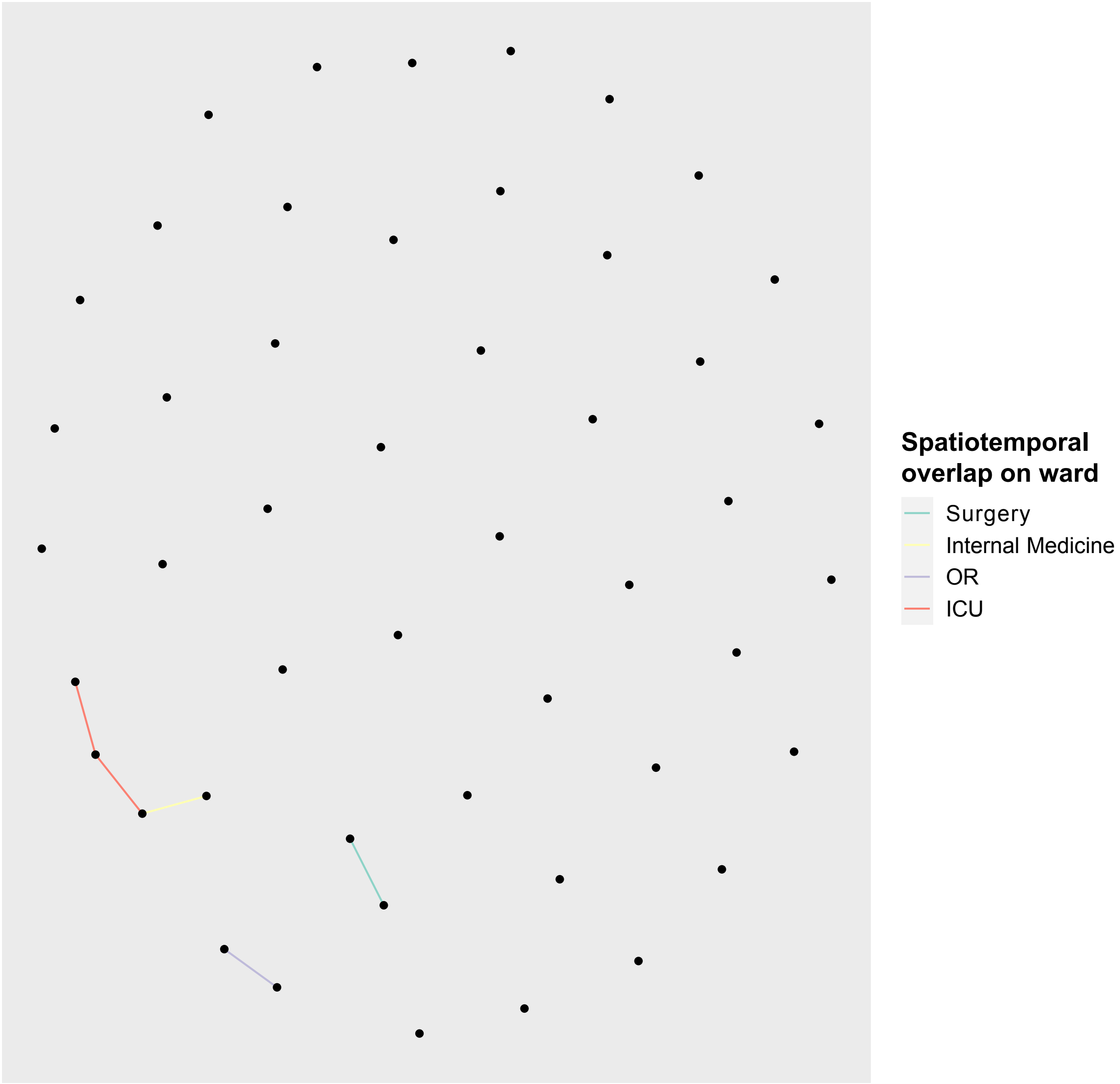


Individual patients are shown as vertices. Edges were drawn if there was spatiotemporal overlap on a hospital wards (indicated by edge colour) and a genomic criterion of presence of $bla_{NDM}$ in the same plasmid (defined as same plasmid group and NDM variant with ≤5 SNVs difference in the backbone).

Abbreviations: ICU – Intensive Care Unit; NDM – New Delhi metallo-beta-lactamase; OR – Operating Room.
